# Genome-wide association study meta-analysis supports association between MUC1 and ectopic pregnancy

**DOI:** 10.1101/2022.10.31.22281750

**Authors:** Natàlia Pujol Gualdo, Estonian Biobank Research Team, Reedik Mägi, Triin Laisk

## Abstract

Ectopic pregnancy is an important cause of maternal morbidity and mortality worldwide. To better understand the genetic risk factors underlying this pregnancy complication, we conduct a GWAS meta-analysis and identify two genome-wide significant loci on chromosomes 1 (rs4971091, p=5.32×10^−9^) and 10 (rs11598956, p=2.41×10^−8^). Follow-up analyses propose MUC1, an epithelial glycoprotein with an important role in barrier function, as the most likely candidate for the association on chromosome 1. We also characterise the phenotypic and genetic correlations with other phenotypes, identifying a genetic correlation with smoking and diseases of the (genito)urinary and gastrointestinal system, and phenotypic correlations with various reproductive health diagnoses, reflecting the previously known epidemiological associations.

## Introduction

An ectopic pregnancy is a pregnancy complication where the fertilized oocyte implants and grows outside the uterine cavity, mostly (95% of cases) in the Fallopian tube. Studies suggest that tubal ectopic pregnancy is a result of a combination of retention of the embryo within the Fallopian tube due to impaired embryo-tubal transport and alterations in the tubal environment allowing premature implantation^1^. Ectopic pregnancy is one of the most common early pregnancy complications, affecting around 1-2% of all pregnancies, and is also the major cause of maternal mortality in the first trimester, accounting for approximately 6% of all maternal deaths^2^.

The symptoms of ectopic pregnancy include abdominal pain and/or vaginal bleeding, and it is diagnosed by human choriogonadotropin (hCG) testing and ultrasound. Although in some cases an ectopic pregnancy miscarries on its own, usual treatment includes methotrexate or surgery.

Known risk factors of ectopic pregnancy include maternal age, smoking, tubal surgery, tubal adhesions or blockage due to pelvic inflammatory disease, and the use of assisted reproductive technologies. Having an ectopic pregnancy also increases the risks of having another one, with a recurrence rate of 5-25%^2^. Women with a previous ectopic pregnancy are also at an increased risk of compromised future fertility, post-traumatic stress, depression, and anxiety^3,4^.

It has been shown that daughters of mothers with ectopic pregnancy have a 50% higher risk of ectopic pregnancy compared to daughters of women without an ectopic pregnancy^5^, indicating a potentially heritable component to the condition. A recent study in mice suggested the gene *Adgrd1* might control oviductal fluid flow and embryo transit and thus be involved in ectopic pregnancy^6^. Similarly, ectopic pregnancies have been reported in individuals with primary ciliary dyskinesia, a condition with genetic origins^7,8^. However, no large-scale studies to explore the potential contribution from genetic factors in ectopic pregnancy have been conducted.

Here, we report a genome-wide association study meta-analysis in 7070 ectopic pregnancy cases and 248,810 controls. We identify two genome-wide significant signals and thus provide the first evidence of a genetic susceptibility component to ectopic pregnancy in humans.

## Methods

### Study cohorts

Our analyses included a total of 7070 women with ectopic pregnancy and 248,810 controls of European ancestry from two studies: summary level statistics from the FinnGen R7 data release (4526 cases and 124,547 controls) for the phenotype O15_PREG_ECTOP, and individual level data from the Estonian Biobank (EstBB, 2544 cases and 124,263 controls). In the EstBB, cases were defined as women having ectopic pregnancy diagnosed by International Classification of Disease (ICD-10) diagnosis code O00. Controls were defined as individuals who did not have the respective ICD code. We ran an additional analysis in the EstBB identifying controls as women who have ever been pregnant or have ever delivered (obtained from questionnaire data or from ICD-codes: Z32 - Pregnancy examination and test, O80-O84 - Delivery), this analysis included 53,671 controls.

### Cohort-level analyses

The EstBB is a population-based biobank with over 200,000 participants (20% of Estonian population), currently including around 135,000 women. All biobank participants have signed a broad informed consent form and analyses were carried out under ethical approval 1.1 - 12/624 from the Estonian Committee on Bioethics and Human Research (Estonian Ministry of Social Affairs) and data release N05 from the EstBB. Individuals with EP were identified using the ICD-10 code O00 (mean age at recruitment =38.4 years, sd=10.5), and all female biobank participants who did not have this diagnosis were considered as controls (mean age at recruitment=44.9 years, sd=16.3), which resulted in 2544 cases and 124,263 female controls. The age range of the selected controls was 49.7+/- 14.2 years. Information on ICD codes is obtained via regular linking with the National Health Insurance Fund and other relevant databases^9^.

Details of EstBB genotyping procedure have been described previously^10–12^. Briefly, all EstBB participants were genotyped using Illumina GSAv1.0, GSAv2.0, and GSAv2.0_EST arrays at the Core Genotyping Lab of the Institute of Genomics, University of Tartu. Individuals were excluded from the analysis if their call-rate was <95% or if their sex defined by heterozygosity of X chromosomes did not match their sex in the phenotype data. Before imputation, variants were filtered by call-rate <95%, HWE *p*-value < 1e−4 (autosomal variants only), and minor allele frequency <1%. Same analyses were conducted for association analysis and imputation of chromosome X, except for the HWE filter, which was not applied. Pre-phasing was conducted using Eagle v2.3 software^13^ and imputation was done using Beagle v.28Sep18.793^14^. The population specific imputation reference of 2297 whole genome sequencing samples was used^15^. Association analysis was carried out using REGENIE (v2.2.4)^16^ with year of birth and 10 PCs as covariates in step I, and by default excluding variants with a minor allele count <5. Analyses were run both using an additive and recessive model (results for the lead signals are shown on Supplementary Figure 1). In EstBB, variants with poor imputation quality (INFO score < 0.4) were excluded from downstream association analysis.

For FinnGen, we used GWAS summary statistics from the R7 data release and thus did not have individual level data. The summary statistics were downloaded as described here: https://www.finngen.fi/en/access_results. The FinnGen cohort and the relevant genotyping/data analysis details have been described in Kurki et al.^17^. Briefly, REGENIE 2.0.2^16^ was used for analysis, with age, 10 PCs, and genotyping batch as covariates. FinnGen summary statistics include variants with a minor allele count >5 and imputation INFO score 0.6.

### GWAS meta-analysis

We conducted an inverse variance weighted fixed-effects meta-analysis with genomic control using GWAMA (v2.1)^18^. The genomic inflation factors (lambda) of the individual study summary statistics were 1.037 (EstBB), and 1.036 (FinnGen). Genome-wide significance was set to p < 5 × 10^−8^ and for downstream analyses we included only variants present in both cohorts (n=12,363,169 variants).

### Annotation of GWAS signals

We used FUMA (v1.4.0)^19^ to annotate the GWAS signals. FUMA is an online platform that performs annotation of GWAS signals using data from several databases. First, FUMA identifies lead SNPs (p-value <5 × 10^−8^ and LD r2<0.1, based on 1000G European reference) and independent significant SNPs and each risk locus (p-value <5 × 10^−8^ and LD r2< 0.6). Then FUMA identifies potential candidate SNPs that are in LD with any of the identified independent significant SNPs and annotates them via linking with several databases (ANNOVAR^20^, RegulomeDB^21^, CADD^22^ scores etc.), which gives information on their location, functional impact, and potential regulatory effects (Supplementary Table 1).

FUMA also links with the GWAS catalog (https://www.ebi.ac.uk/gwas/) for previous associations between the identified candidate SNPs and studies in the GWAS catalog. The results of this look-up are presented in Supplementary Table 2.

### Colocalisation analysis

We used HyPrColoc^23^, a colocalisation method for identifying the overlap between our GWAS meta-analysis signals and cis-QTL signals from different tissues and cell types (expression QTLs, transcript QTLs, exon QTLs and exon usage QTLs available in the eQTL Catalogue^24^. We lifted the GWAS summary statistics over to hg38 build to match the eQTL Catalogue using binary liftOver tool (https://genome.sph.umich.edu/wiki/LiftOver#Binary_liftOver_tool).

For the genome-wide significant (p<5 × 10^−8^) GWAS loci identified we extracted the +/-500kb of its top hit from QTL datasets and ran the colocalization analysis against eQTL Catalogue traits. For each eQTL Catalogue dataset we included all the QTL features which shared at least 80% of tested variants with the variants present in our GWAS region. We used the default settings for HyPrColoc analyses and did not specify any sample overlap argument, because HyPrColoc paper^23^ demonstrates that assuming trait independence gives reasonable results. HyPrColoc outputs the posterior probability that genetic association signals for those traits are colocalising (we considered two or more signals to colocalize if the posterior probability for a shared causal variant (PP4) was 0.8 or higher. All results with a PP4 > 0.8 can be found in Supplementary Table 3).

### Look-up of main signals in Biobank Japan

We looked up the association results for our European ancestry meta-analysis lead variants with summary statistics publicly available from Biobank Japan^25^, including 605 cases and 82,156 controls to compare association results in a different ancestry cohort in association with ectopic pregnancy. The Biobank Japan data was downloaded from https://pheweb.jp.

### Heritability analysis

We used LD-Score (LDSC) regression^26^ and the HapMap3 reference panel to estimate the total SNP-based heritability (*h*^2^_SNP_) of the ectopic pregnancy meta-analysis. We assumed a population prevalence of 1% and a study sample prevalence of 2%.

### Genetic correlation analysis

We used the Complex Traits Genetics Virtual Lab (CTG-VL, https://genoma.io/) to calculate genetic correlations between our ectopic pregnancy meta-analysis and 1335 traits. We applied a multiple testing correction (FDR < 5%) to determine statistical significance using the p.adjust function in R 3.6.3. Results of the genetic correlation analysis are presented in Supplementary Table 4 and Figure 2.

### Gene-based analysis and look-up of previous candidate genes

Gene-based testing was carried out with MAGMA v1.08 implemented in FUMA^19,27^. Additionally, we queried previous proposed candidate genes for ectopic pregnancy amongst our summary statistics results: *Adgrd1* (also known as *GPR113*)^6^, *VEGFA* (vascular endothelial growth factor A), *IL8* (interleukin 8), *IL6* (interleukin 6), *ESR1* (estrogen receptor 1) and *EGFR* (epidermal growth factor receptor)^28^. Results of this analysis can be found in Supplementary Table 5.

### Analysis of associated phenotypes in EstBB

Using the individual level data in the EstBB, we conducted an analysis to find ICD10 diagnosis codes associated with the O00 diagnosis. We tested the association between ectopic pregnancy (defined as ICD10 O00) and other ICD10 codes in a logistic regression framework, adjusting for age and 10 genetic PCs. Since the EstBB includes a large proportion of relatives and inclusion of relatives might inflate the association statistics, we excluded all first- and second-degree relatives (pi-hat cut-off value 0.2) in pairwise comparisons, keeping the index cases, if possible, to not lose in power. This resulted in 2370 cases and 42,970 controls for the analysis Bonferroni correction was applied to select statistically significant associations (number of tested ICD main codes – 2001, corrected p-value threshold – 2.5 x10^−5^). Results were visualised using the PheWas library 0.99.5-4. All analyses were carried out in R 3.6.3. The results of this analysis are presented in Supplementary Table 6. We carried out the analysis using both unselected controls (all women) and selected controls (women who have been pregnant but have not had ectopic pregnancy). For clarity, we only present the results from the selected controls analysis.

## Results

### Genome-wide association study meta-analysis

The meta-analysis identified two loci for ectopic pregnancy, with three independent lead signals significantly associated with EP (*P* < 5 × 10−8). There was no evidence of inflation (λ = 1.0285) in the GWAS meta-analysis (LDSC intercept= 0.9706 (s.e. 0.0067)). The observed SNP heritability estimate was 0.0106 (s.e. 0.0019), which corresponds to a liability scale SNP heritability of 7.4 % (s.e. 0.13).

The first signal is a common variant (minor allele frequency 0.45) on chromosome 1 (lead signal rs4971091, p=5.32×10^−9^), in the exon of a non-coding transcript *KRTCAP2* (Table 1). According to FUMA, there is another independent SNP rs1057941 (p=1.38×10^−8^) in the GBAP1 pseudogene. As can be seen from Figure 1, the signal is in a gene-dense region that also includes *MUC1*. Colocalisation analysis showed our GWAS signal overlaps with QTL signals associated with both *MUC1* expression and specific transcripts (Supplementary Table 3). Further analysis of the association signal revealed that the second independent SNP in this region, rs1057941, is in LD with rs4072037 (r2_EUR=0.52, r2_FIN=0.76, r2_EST=0.69), a splice acceptor variant in the 2nd exon of *MUC1* that in our meta-analysis has a p-value of 1.87 × 10^−7^ (C-allele OR=1.10 95% CI 1.06-1.13). According to the GWAS catalog look-up, this region has previously been associated with several biomarker levels, but also with gastric cancer (Supplementary Table 2).

**Table 1.**
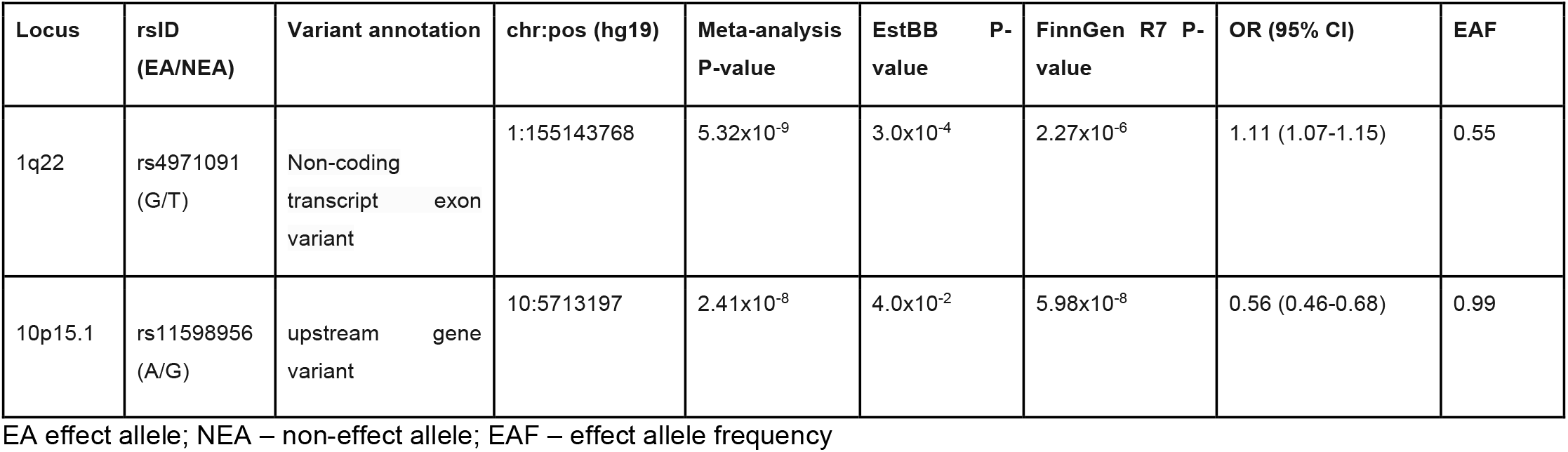
Genome-wide significant lead variants in the two loci associated with ectopic pregnancy identified in the GWAS meta-analysis. Positions are according to build GRCh37.

**Figure 1.**
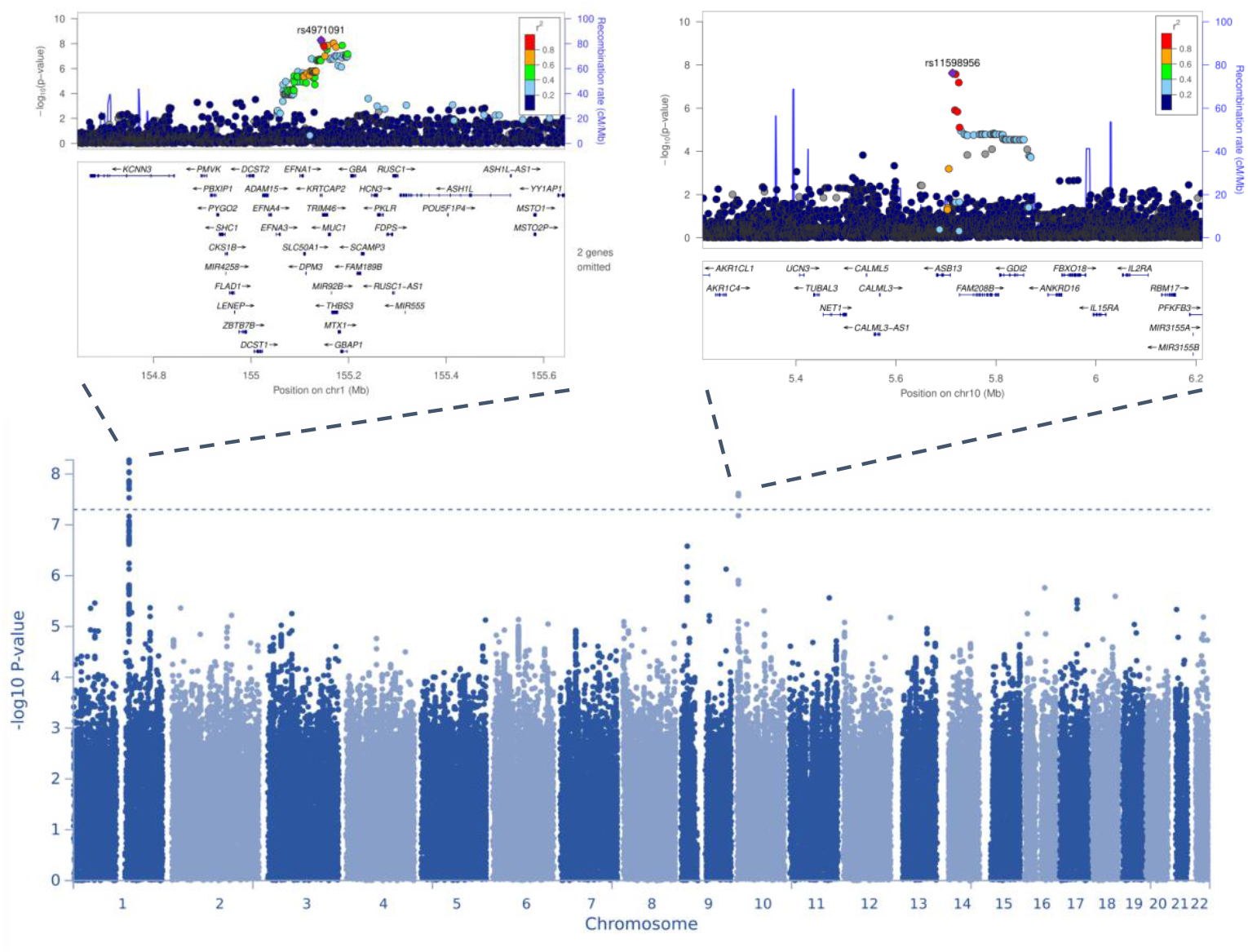
Manhattan and locus zoom plots for GWAS meta-analysis for ectopic pregnancy. On the Manhattan plot, the *y* axis represents −log_10_(*P-*values) for association of variants with ectopic pregnancy. The horizontal dashed line represents the threshold for genome-wide significance (*P* < 5 × 10^−8^). Regional plots display the lead variants of genome-wide significant loci on chromosomes 1 (left) and 10 (right), respectively.

**Figure 2.**
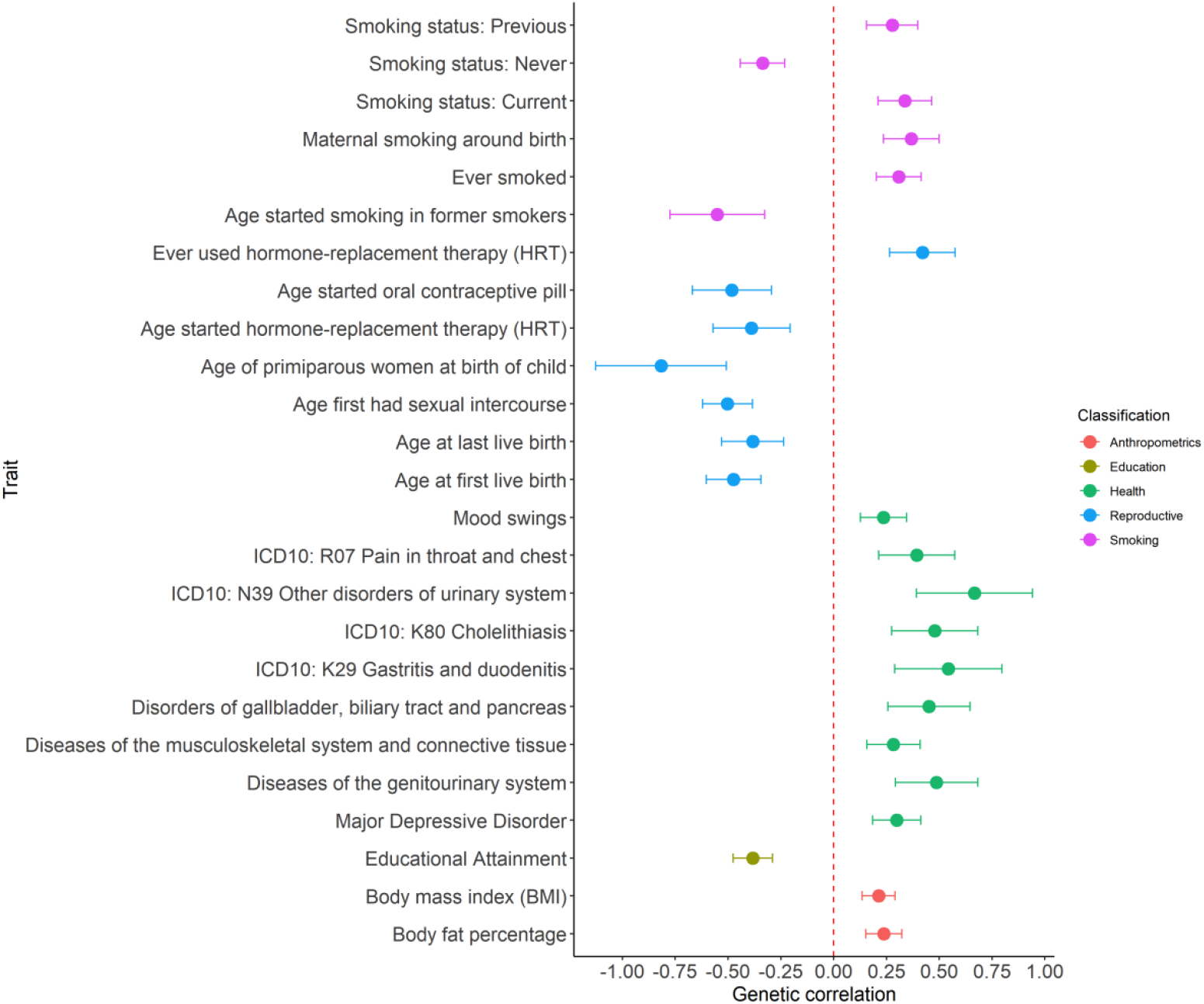
Genetic correlations between ectopic pregnancy and anthropometric, educational, health-related, reproductive, and smoking phenotypes. Center values show the estimated genetic correlation (rg), which is presented as a dot and error bars indicate 95% confidence limits. Dotted red line indicates no genetic correlation.

We identified a second signal on chromosome 10, where the lead SNP is rs11598956 (p=2.41×10^−8^, minor allele frequency 0.6%, imputation INFO score >0.9 in both cohorts) between the *ASB13* and *TASOR2* genes. Of the candidate SNPs at this locus, rs74599685 in the intron of *ASB13* has a RegulomeDB score of 2b, which indicates evidence for a regulatory effect.

A sensitivity analysis carried out in the EstBB using controls who have been pregnant yielded similar results (data not shown), indicating that the use of unselected controls does not have an effect on the results.

Look-up of lead variants in the Biobank Japan dataset consisting of 605 cases and 82,156 controls showed that although neither of the variants was statistically significantly associated with ectopic pregnancy, the effect directions were the same as in the European ancestry meta-analysis (rs4971091 G allele OR=1.12, 95% CI 0.97-1.31; rs11598956 G allele OR=1.07 95% CI 0.79-1.46)

### Gene-based analysis and look-up of previous candidate genes with MAGMA

Gene-based analysis implemented in MAGMA identified six associations significant after Bonferroni correction (0.05/18895 protein coding genes), all on chromosome 1 - *RP11-201K10*.*3, THBS3, KRTCAP2, TRIM46, MTX1*, and *EFNA1*. Look-up of genes associated with ectopic pregnancy in previous literature (*ADGRD1* (*GPR113*), *VEGFA* (vascular endothelial growth factor A), *IL8* (interleukin 8), *IL6* (interleukin 6), *ESR1* (estrogen receptor 1) and *EGFR* (epidermal growth factor receptor) revealed that none of them passed the Bonferroni correction threshold and only *GPR113* (also known as *ADGRD1*) and *ESR1* were nominally significant (p=0.046 and p=0.044, respectively). Full results are shown in Supplementary Table 5.

### Genetic correlation

We tested pairwise genetic correlations (*r*_g_) between ectopic pregnancy and 1335 other traits (Supplementary Table 4) available from the Complex Traits Genetics Virtual Lab (CTG-VL, https://genoma.io/). We found 93 significant (FDR < 0.05) genetic correlations with European-ancestry ectopic pregnancy meta-analysis. Selected genetic correlations with phenotypes related to smoking, overall and reproductive health, anthropometrics and education are presented in Figure 2. As expected, risk of ectopic pregnancy is associated with different smoking phenotypes. We also observed positive genetic correlations with diseases of the (genito)urinary and gastrointestinal system.

### Associated phenotypes

The significantly associated disease codes in the pheWAS analysis are consistent with what is known about the etiopathogenesis of the condition, validating the used phenotype definition. In our analysis, women with a diagnosis of ectopic pregnancy have significantly more diagnoses of female infertility (N97, OR=3.4 (3.1-3.7)), procreative management (including IVF, Z31, OR=3.3 (2.9-3.7)), salpingitis (N70, OR=2.6 (2.3-2.9)), posthemorrhage anemia (D62, OR=6.3(5.0-7.9)), miscarriage (O02, OR=2.3 (2.0-2.5); O03, OR=2.6 (2.3-3.0)), other female pelvic inflammatory disease (N73, OR=3.2 (2.7-3.7)), chlamydial infection (A56, OR=1.6 (1.4-1.9)), bleeding in early pregnancy (O20, OR=2.1 (1.9-2.3)), abdominal pain (R10, OR=1.6 (1.5-1.8)). In addition, we found increased odds of peritoneal disorders (K66, OR=5.7 (4.4-7.3)), habitual abortion (N96, OR=3.3 (2.7-4.1)), excessive, frequent and irregular menstruation (N92, OR=1.5 (1.4-1.7)), complications associated with artificial insemination (N98, OR=4.4 (3.2-5.9)), ovarian dysfunction (E28, OR=1.5 (1.4-1.7)), endometriosis (N80, OR=1.7 (1.5-2.0)), hydatidiform mole (O01, OR=6.0 (3.8-9.5)) and three ICD codes related to the respiratory system - acute nasopharyngitis (J00), acute tonsillitis (J03), and acute bronchitis (J20). In total, 61 diagnosis codes with a significantly different prevalence in cases and controls were identified (Figure 3, Supplementary Table 6)

**Figure 3.**
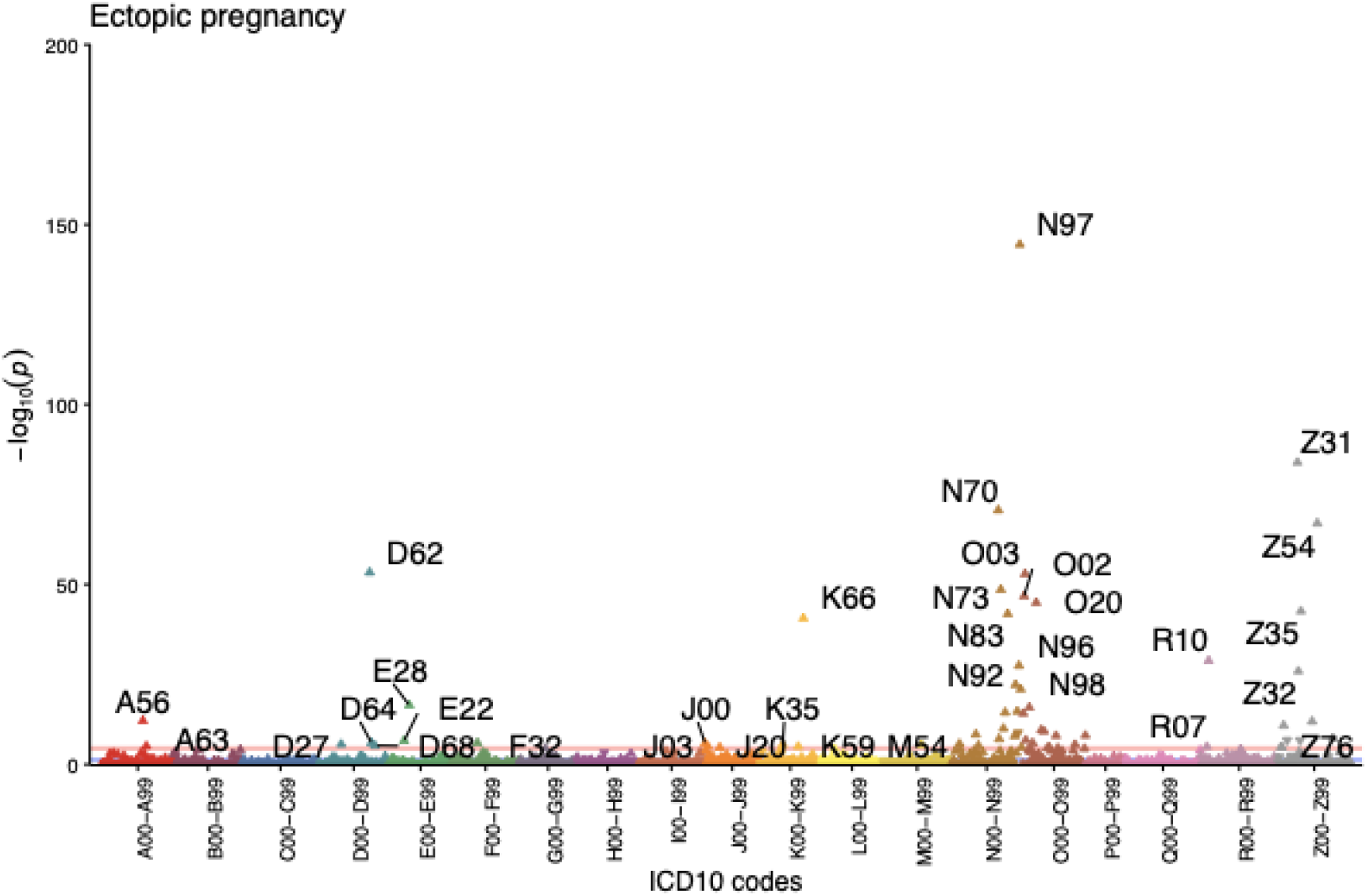
Disease codes associated with a diagnosis of ectopic pregnancy (O00) in the Estonian Biobank. Each triangle represents one ICD-10 maincode, while different colors correspond to different chapters. The direction of the triangle illustrates effect direction - upward pointing triangles show increased prevalence of the diagnosis code in ectopic pregnancy cases. The pink line shows the Bonferroni corrected threshold for statistical significance.

## Discussion

We conducted the first GWAS meta-analysis for ectopic pregnancy in 7070 women with ectopic pregnancy and 248,810 controls from two European-ancestry biobanks. We identify two genetic risk loci providing evidence of a genetic susceptibility component to this common early pregnancy complication. Our results inform the genetic background and provide clues to the etiopathogenesis of ectopic pregnancy.

We also characterise the phenotypic and genetic correlations with other phenotypes. The genetic correlation analyses provide further support for smoking as an important risk factor of ectopic pregnancy. A positive correlation with maternal smoking around birth may point towards a transgenerational effect on tubal function, similar to the one observed in the respiratory tract of offspring of smoking mothers^29^. However, caution is needed when interpreting this result since as far as we are aware, this is the first report of such an association. Phenotype-level analyses show increased prevalence of various reproductive health diagnoses in women with ectopic pregnancy, reflecting the previously known epidemiological associations between ectopic pregnancy and IVF, salpingitis, pelvic inflammatory disease, and others, and lending support to our ICD-code based phenotype definitions. While to some extent the increased prevalence of these disease codes in women with ectopic pregnancy is due to shared socio-economic and behavioral risk factors, it is unclear whether in some cases shared molecular mechanisms might also play a role. We also observe an increased prevalence of respiratory diagnoses related to sinusitis, tonsillitis, and bronchitis, which can all be linked to altered ciliary function, similar to ectopic pregnancy, since cilia in the respiratory tract epithelium help to clear mucus and protect against infections^30^.

Recent years have seen numerous studies that have begun to clarify the genetic susceptibility to pregnancy complications such as pre-eclampsia^31^ or gestational diabetes^32^. Less is known about the genetics of early pregnancy complications, which often are symptoms of unviable pregnancy and thus inevitably have a lower heritability. By combining data from two large population-based biobanks, we were able to reach a sample size that allowed us to identify the first genetic susceptibility factors for ectopic pregnancy and demonstrate a heritable component to the condition. Our research underlines the value of large-scale biobank resources to advance the study of pregnancy complications, of remarkable importance since these lack suitable animal models and there are ethical concerns on the use of human biopsies.

Our study provides genetic evidence supporting the role of *MUC1* in ectopic pregnancy. *MUC1* is a large epithelial apical surface glycoprotein that acts as a barrier to embryo implantation. Previous small-scale studies have shown reduced expression of MUC1 in the Fallopian tubes of women diagnosed with ectopic pregnancy^33–35^. This has led to the hypothesis that altered expression of *MUC1* may predispose women to ectopic pregnancy. The results of the current study support this hypothesis, as the GWAS signal on chromosome 1 colocalises with quantitative trait loci affecting both *MUC1* gene expression and specific transcript expression. Moreover, we were able to tie our association with a splice acceptor variant rs4072037, providing insight to the potential functional mechanisms underlying this association.

The T- allele (in previous literature referred to as the A allele) of rs4072037 results in a 27bp/9 amino acid deletion in the 2nd exon of MUC1. Previous studies have shown that this may lead to changes in intracellular trafficking, glycosylation, and folding of the protein, which all may affect the function of MUC1, or alternatively, it has been shown that the same variant may also affect the transcriptional activity of the *MUC1* promoter^36^. This variant is first and foremost known for its association with gastric cancer^36–38^, and in this context, it is believed rs4072037 influences the quantity and/or the quality of the MUC1 protein. This causes a difference in its barrier function in the stomach and subsequently modifies the susceptibility to environmental risk factors that cause inflammation and carcinogenesis. Our genetic correlation results showing association between ectopic pregnancy and gastrointestinal diagnoses also reflect the shared genetic background between these two conditions. Altered barrier function of MUC1 in the Fallopian tubes is also a plausible explanation for its association with ectopic pregnancy. In line with this, rs4072037 has been associated with pregnancy loss and ectopic pregnancy also in the UK Biobank^39^. However, due to the age structure of the UK Biobank, the results should be interpreted with caution since many of the actual cases are mislabelled as controls, resulting in extremely low prevalence of pregnancy phenotypes in this biobank cohort.

The association with the second locus on chromosome 10 is less clear and our GWAS follow-up analyses failed to provide any solid evidence to support the role of any specific gene. Given the low frequency of this variant (minor allele frequency <1%), further replication studies are needed in populations where it is more common to confirm its association with ectopic pregnancy and propose potential explanations about the mechanisms.

While our study of more than 7000 cases identifies two genetic risk loci for ectopic pregnancy, further larger meta-analyses or independent studies are needed to validate these findings, especially regarding the rare variant on chromosome 10. Moreover, as with other reproductive phenotypes, the lack of sufficiently sized relevant tissue data (in this case, Fallopian tube) in commonly used gene expression and other databases (such as the GTeX), hinders the proper interpretation of GWAS findings and highlights the need for large-scale gene expression studies in female reproductive tissues. Additional studies are also needed to evaluate whether smoking, an important risk factor of ectopic pregnancy, somehow mediates the genetic effects, along with studies evaluating additional risk factors and their interaction with ectopic pregnancy.

In conclusion, the first genome-wide association study meta-analysis in ectopic pregnancy provides genetic evidence to support the involvement of the MUC1 epithelial glycoprotein and maps genetic and phenotypic associations with other phenotypes, providing input for further studies.

## Data Availability

All data produced in the present study will be made available once the manuscript has been published.

## Acknowledgements

NPG was supported by MATER Marie Sklodowska-Curie which received funding from the European Union’s Horizon 2020 research and innovation program under grant agreement No. 813707. This study was funded by European Union through the European Regional Development Fund Project No. 2014-2020.4.01.15-0012 GENTRANSMED. Computations were performed in the High-Performance Computing Center of University of Tartu. We want to acknowledge the participants of the Estonian Biobank, and participants and investigators of the FinnGen study. The writing of this paper was supported by the writing retreats organised by the Institute of Genomics, University of Tartu.

